# How do perceptions of Covid-19 risk impact pregnancy-related health decisions? A convergent parallel mixed-methods study protocol

**DOI:** 10.1101/2023.07.10.23292463

**Authors:** Meredith Vanstone, Rebecca H. Correia, Michelle Howard, Elizabeth Darling, Hamideh Bayrampour, Andrea Carruthers, Amie Davis, Dima Hadid, Erin Hetherington, Aaron Jones, Sujane Kandasamy, Cassandra Kuyvenhoven, Jessica Liauw, Sarah D. McDonald, Caroline Mniszak, Monica L. Molinaro, Manisha Pahwa, Tejal Patel, Marina Sadik, Njideka Sanya, Katrina Shen, Devon Greyson

**Author notes:** Correspondence: M Vanstone, Department of Family Medicine. McMaster University, 100 Main St W Room 5001F, Hamilton, Ontario Canada, L8P 1H6. Registration: Clinicaltrials.gov registration number: NCT05663762.

## Abstract

**Introduction:** Pregnant people have a higher risk of severe COVID-19 disease. They have been disproportionately impacted by COVID-19 infection control policies, which exacerbated conditions resulting in intimate partner violence, healthcare access, and mental health distress. This project examines the impact of accumulated individual health decisions and describes how perinatal care and health outcomes changed during the COVID-19 pandemic.

**Objectives:**

1. *Quantitative strand:* Describe differences between 2019, 2021, and 2022 birth groups related to maternal vaccination, perinatal care, and mental health care. Examine the differential impacts on racialized and low-income pregnant people.
2. *Qualitative strand:* Understand how pregnant people’s perceptions of COVID-19 risk influenced their decision-making about vaccination, perinatal care, social support, and mental health.

**Methods and analysis:** This is a Canadian convergent parallel mixed-methods study. The <u>quantitative strand</u> uses a retrospective cohort design to assess birth group differences in rates of Tdap and COVID-19 vaccination, gestational diabetes screening, length of post-partum hospital stay, and onset of depression, anxiety, and adjustment disorder, using administrative data from ICES, formerly the Institute for Clinical Evaluative Sciences (Ontario) and PopulationData BC (PopData) (British Columbia). Differences by socioeconomic and ethnocultural status will also be examined. The <u>qualitative strand</u> employs qualitative description to interview people who gave birth between May 2020-December 2021 about their COVID-19 risk perception and health decision-making process. Data integration will occur during design and interpretation.

**Ethics and dissemination:** This study received ethical approval from McMaster University and the University of British Columbia. Findings will be disseminated via manuscripts, presentations, and patient-facing infographics.

**Strengths and limitations of this study:**

- Population-based administrative data cohorts are very large, ensuring that analyses are high-powered.
- Mixed-methods design will allow us to offer explanation for changes in healthcare use observed through administrative data.
- Cross-provincial design permits examination of the potential impacts of COVID-19 infection prevention and control policies on pregnant people’s health.
- Use of Canadian Index of Multiple Deprivation will allow us to examine differences in healthcare use according to economic, racial, and immigration factors.
- Team includes 5 co-investigators with lived experience of pandemic pregnancies.

## Introduction

Pregnant people experience higher rates of COVID-19 hospitalization, intubation, and death compared to similar age and sex-matched peers who are not pregnant.^1-3^ As in the general population, pregnant people who are racialized, low-income, or who live in high-density neighbourhoods have elevated COVID-19 infection rates.^3 4^

While there is little evidence specific to pregnant people, evidence about health care access and well-being in women, trans, and gender non-binary people helps us understand the circumstances faced in pandemic pregnancies. Most pregnant people identify as women, and women have been disproportionately impacted by the COVID-19 pandemic.^5 6^ Overall well-being of pregnant people has also been jeopardized by COVID-19 social isolation policies resulting in increased intimate partner violence ^7-9^ and mental health distress.^10 11^ During the pandemic, women experienced unemployment and income loss partly attributed to precarious employment and increased childcare responsibilities.^12 13^ Women were also overrepresented in designated essential service jobs with high levels of COVID-19 exposure (e.g., nursing and patient service occupations).^14^ Trans and non-binary people faced disproportionate pandemic-related challenges to mental health, violence, income loss, and access to healthcare compared to people of other genders.^15 16^

This project examines pregnant people’s health decisions within workplace, home, and community environments, describing their accumulated impact on key pregnancy outcomes and care indicators related to three themes: 1) vaccination, 2) perinatal care, social supports and mental health. The decisions made during pregnancy have longitudinal impacts on the life of the pregnant person, future child, and family.^17^ Given the likely unique and elevated health, social, and economic challenges faced by pregnant people during the pandemic, and the importance of health decisions during pregnancy, it is necessary to understand how pandemic circumstances have shaped decisions about accessing pregnancy-related healthcare. Understanding how and why pregnant people make health decisions allows for targeted clinical and social support as the pandemic endures and will inform future policy planning.

### Theme 1: Vaccination

Although pregnant people were not deliberately included in pre-market clinical trials, based on international immunization registry data ^18^, in May 2021 the Canadian National Advisory Committee on Immunization (NACI) recommended COVID-19 vaccination for pregnant people, with a preference for mRNA vaccines. Accordingly, pregnant populations were prioritized for access in late April (Ontario) and early May (British Columbia) 2021.^19 20^ COVID-19 mRNA vaccines have shown a good safety profile during pregnancy ^21^, and protect against severe disease in the pregnant person as well as negative neonatal outcomes ^22^ including stillbirth ^23^ and infant hospitalization for COVID-^24^

Since 2018, Canadian guidelines have recommended that all pregnant people be offered pertussis vaccination (available via combination tetanus toxoid, diphtheria, and acellular pertussis, or Tdap, vaccine).^25 26^ Uptake of routine (Tdap, influenza) vaccination during pregnancy in Canada has lagged behind that seen in comparator countries, but increased during the COVID-19 pandemic^27^. However, this overall increase obscures growing disparities (e.g., by ethnicity, income) and access barriers that may have been exacerbated by the pandemic^27^. Further, there have been indications of greater vaccination delay or refusal during pregnancy than in the general population, largely due to safety concerns.^28 29^ Factors commonly associated with low uptake of vaccination during pregnancy include socioeconomic status, demographic and geographic characteristics, access to prenatal care, and health literacy ^30-32^. While a recommendation from a trusted prenatal care provider is typically understood to be one of the strongest influences for vaccine uptake ^32-36^, limited in-person appointments may have weakened that influence during the pandemic. We do not yet know how vaccine uptake has changed, including how decreased access to in-person prenatal care may have influenced decisions regarding vaccines recommended in pregnancy.

### Theme 2: Perinatal Care

Perinatal care includes medical care delivered during the prenatal, intrapartum and postpartum periods. Routine prenatal care consists of a series of regular interactions with a health care provider – typically a family physician, midwife, and/or obstetrician (OB) – for education, screening, and treatment interventions.^37^ Inadequate prenatal care has been associated with a number of negative health outcomes, including stillbirth, pre-term birth, low birth weight, NICU admission, and postpartum depression and anxiety.^38^ The COVID-19 pandemic instigated many changes to the delivery of routine prenatal care related to the incorporation of telephone or virtual visits,^39-41^ and changes to prenatal care schedules to adapt to re-allocation of healthcare resources, personal protective equipment limitations, and lack of continuity of care from the same prenatal care provider.^42^ Perinatal care also changed in relation to policies that reduced support persons and visitors, and exacerbated other access issues related to childcare, precarious work, and transportation.^43-45^ There are little data about the outcomes associated with these changes to prenatal care delivery, with some studies finding no difference in maternal and infant health outcomes ^46 47^ and others in a global meta-analysis demonstrating shorter in-hospital stays after delivery, less prenatal care attendance, lower participation in screening for infection, anemia and fetal anomaly.^47^ We do not know how these changes to the delivery of prenatal care influenced features such as uptake of gestational diabetes screening, or the duration of postpartum hospitalization in Canada. Further, it is unknown whether any changes in healthcare access that may have occurred reflected patient choice or influenced patient experience.

### Theme 3: Mental Health and Social Support

Untreated antenatal depression and anxiety are associated with pre-term birth, low birth weight, and increased risk of infant hospitalization and death within the first year of life. ^48 49^ A systematic review and meta-analysis of global data indicate high rates of prenatal depression (30.5%) and anxiety (25.6%) during the COVID-19 pandemic.^50^ Pregnant people in Canada may experience even higher rates of depression and anxiety,^51^ perhaps in response to concerns about threats of COVID-19 to themselves or their fetus, insufficient prenatal care, strained relationships, and social isolation.^52^ Mental health concerns may be greater for those who experienced extra stress or safety fears due to income disruptions, difficulties balancing childcare and work responsibilities, are single parents, or had difficulty obtaining childcare.^11 53,54^ The stress and anxiety experienced during the pandemic may have contributed to lower birth weights, younger gestational age at birth, and other birth-related issues.^42 52^ So far, there are little data about how perinatal mental health diagnosis rates have changed throughout the pandemic. We do not know whether any observed changes in mental healthcare use reflect barriers to accessing care. We do not know how pregnant people experienced their feelings of depression and anxiety, and how they chose to cope with these experiences.

#### Objectives, questions and hypotheses

This research study has two objectives:

1. Evaluate potential associations between exposure(s) and outcome(s) for births occurring in 2019 (Jan 1 2019 – Mar 31 2019 births), 2021 (Jan 1 2021 – Mar 31 2021 births), and 2022 (Jan 1 2022-Mar 31 2022 births) birth groups in Ontario and BC related to vaccination, perinatal care, and mental health, overall, by race/ethnicity and by income. *(Quantitative strand)*
2. Understand how people who gave birth in Ontario or BC in 2020 or 2021 perceived COVID-19 risk and how pandemic circumstances influenced their decision-making about key elements of pregnancy, including vaccination, perinatal care, social support and mental health. *(Qualitative strand)*

Research questions and hypotheses have been operationalized according to our three themes:

### Theme 1: Vaccination

- Quantitative Research Question (RQ): Were rates of vaccination different between 2019, 2021, and 2022 birth groups? What factors are associated with vaccination rates? **Outcomes:** Tdap (ON) and COVID-19 (ON and BC) vaccination rates
  - Hypothesis 1.1: 2019 group will have a higher rate of Tdap vaccination than the 2021 and 2022 groups.
  - Hypothesis 1.2: COVID-19 vaccination rates in the 2022 group will be lower than those of the comparable overall female population in each province.
  - Hypothesis 1.3: In the 2022 group, pregnant persons who receive Tdap vaccination will be more likely to receive COVID-19 vaccination than those who do not receive Tdap vaccination. (Ontario only due to data availability)
- Qualitative RQ: How have pregnant people’s choices about Tdap and COVID-19 vaccination been influenced by perceptions of COVID-19 risk, COVID-19 policies and personal socioeconomic circumstances during the pandemic (“pandemic circumstances”)?
  - How do personal identities and circumstances (e.g., race, education, age, income, presence of other children in the home) affect these decision-making processes?

### Theme 2: Perinatal Care

- Quantitative RQ: Were rates of in-person perinatal care different between 2019, 2021, and 2022 pandemic groups? What factors are associated with in-person perinatal care? **Outcomes:** Gestational diabetes screening, maternal post-partum length-of-stay (BC and ON)
  - Hypothesis 2.1: 2021 group will have lower rates of gestational diabetes screening than the 2019 or 2022 groups.
  - Hypothesis 2.2: 2019 group will have longer length of hospital stay after delivery than 2021 or 2022 groups.
- Qualitative RQ: How have pregnant people’s decisions about seeking prenatal care and planning for birth been influenced by perceptions of COVID-19 risk?
  - How do personal identities and circumstances (e.g., race, education, partnered status, presence of other children in the home) affect risk perception and decision-making processes?

### Theme 3: Mental Health and Social Support

- Quantitative RQ: Were the rates of clinical diagnosis for new depression, anxiety, and adjustment disorders during pregnancy and up to six months postpartum different between 2019, 2021, and 2022 birth groups? What factors are associated with this clinical diagnosis? **Outcomes:** New onset of depression, anxiety and/or adjustment disorder diagnosis during pregnancy and six months postpartum (ON and BC) validated by Frey and colleagues.^55^
  - Quantitative hypothesis 3.1: 2019 group will have lower rates of new clinical diagnosis for depression, anxiety, or adjustment disorder than 2021 and 2022 groups.
- Qualitative RQ: How do pregnant people describe the relationship between their perceptions of COVID-19 risk, experiences of mental health, and decisions about seeking social support during pregnancy?
- Qualitative RQ: How do pregnant people describe their decisions to balance COVID-19 protocols (e.g., masking, limiting social contact) with their physical, mental, and social needs? How do personal identities and circumstances (e.g. race, education, household composition) affect these decision-making processes?

## Methods and Analysis

This project is a parallel mixed-methods study of two Canadian provinces, with thematic data integration at the design and interpretation stages. Ontario and BC were chosen as the two provinces of study because they are populous provinces in Canada with large numbers of live births each year, widespread COVID-19 throughout the pandemic, and access to comprehensive administrative health data relevant for the quantitative arm of this study.

This study was funded in late February 2022. The quantitative cohort creation plans and data access requests were finalized in late Fall 2022. Qualitative data collection was piloted in Summer 2022, and preliminary data from a small sample of Ontario participants was collected in Fall 2022. Qualitative data collection in both provinces is currently ongoing. Study completion is anticipated for February 2024.

### Patient Partnership

Our team includes multiple individuals who have lived experience of pandemic pregnancy. Three co-investigators with lived experience helped to design the study, secure funding, and determine priorities. They contributed to the design of the interview guide and acted as pilot interview participants for interview refinement. Two additional researchers with lived experience conducted interviews and will participate in analysis.

### Quantitative Design and Methods

#### Study design

Using a population-based retrospective cohort design, we will assess differences in rates of clinical care and health outcomes for key pregnancy-related outcomes across three birth groups. A cohort study is appropriate given our interests in making inferences about individuals’ health decision-making and examining individual-level social and clinical exposures. This observational design will enable us to examine the real-world effect of the COVID-19 pandemic on pregnancy-related outcomes within entire jurisdictions .^56^

#### Data sources

Administrative health data from two provinces will be obtained from ICES in Ontario and PopData in British Columbia. These databases are comprehensive sources of health administrative data for healthcare encounters using provincial health insurance in each province. We will link multiple maternal perinatal care datasets at ICES and PopData BC concerning using unique encoded identifiers (person-level deterministic linkage) (Appendix 1).

#### Cohort creation

Our cohort will be defined using an ICES derived cohort (MOMBABY) in Ontario and the BC Perinatal Data Registry, both of which contain medical records for delivering mothers and newborns. Appendix 1 provides details.

#### Study population

Our cohort for the quantitative portion will be defined by three groups according to the date they gave birth: January 1 to March 31, 2019; January 1 to March 31, 2021; and January 1 to March 31, 2022 (Figure 1). These groups represent gestation before the pandemic, gestation during the pandemic but before widespread vaccine availability, and gestation after widespread vaccine availability. These timeframes reflect an assumption that in the experience of pregnant people in Ontario and BC, the pandemic “started” with the enactment of public health measures in mid-March 2020. While the three-month birthing window excludes large portions of the year, this approach is necessary to construct groups within each time period that have no overlap in gestational periods. Overlapping gestational periods would result in cohort members being exposed to multiple time periods, leading to bias in our results.. These groups are standardized by birth month to reflect evidence of seasonality in birth rates and health outcomes.^57 58^

**Figure 1:**
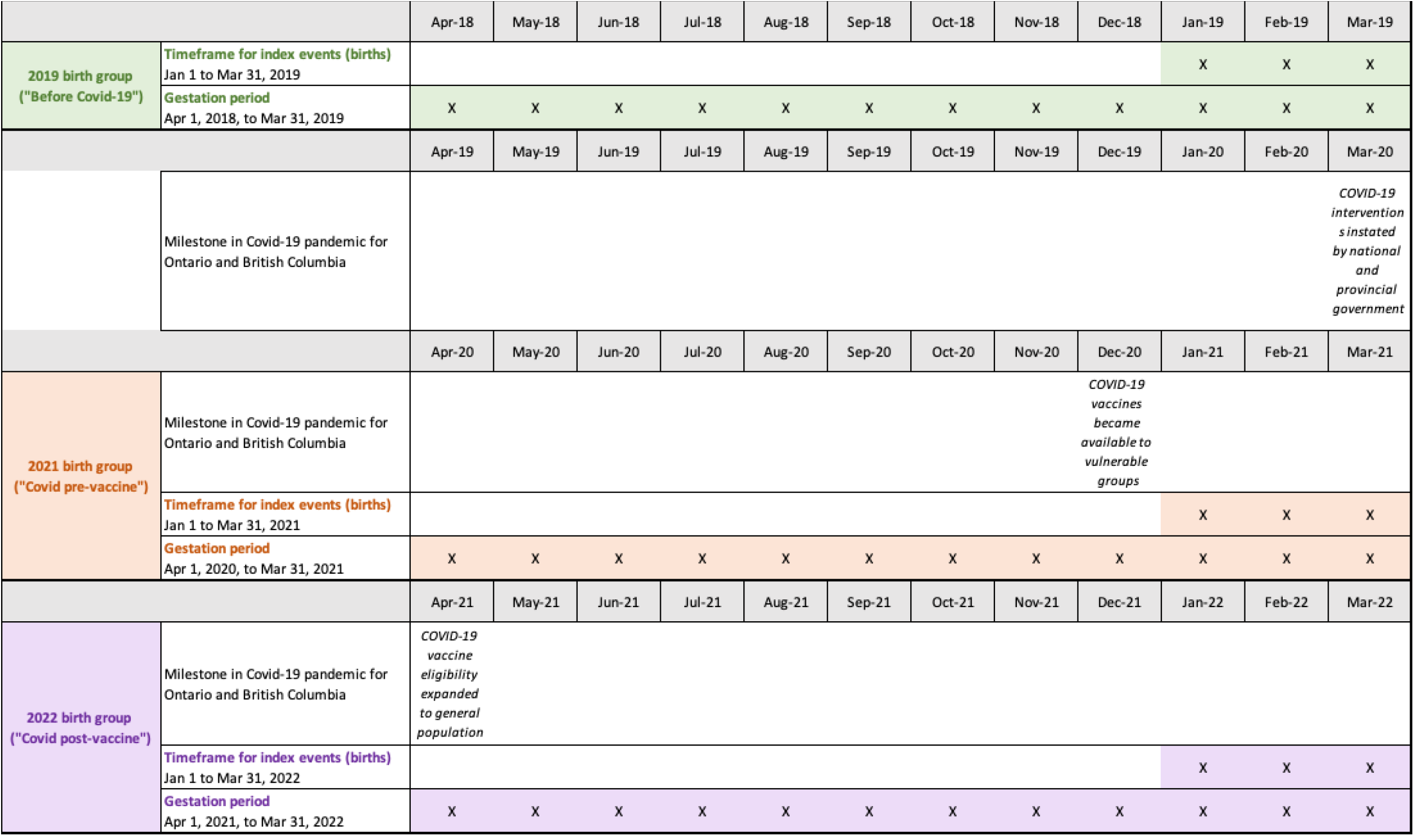
Study population for quantitative portion by birth group

To be eligible for inclusion in our cohort, the individual must have had a live, in-hospital birth during our timeframes of interest. Stillbirths are excluded due to confounding factors around length of stay and mental health. Additional criteria are that the birthing person and newborn must have a valid ICES Key Number (IKN), delivery and birth date, the birthing person must be of female sex, and have been eligible for the provincial health insurance plans in Ontario or BC for the entirety of their perinatal period.

We will also describe pertinent characteristics of linked newborns to mothers in our cohort within MOMBABY and the BC Perinatal Data Registry. In addition, we will create a separate matched cohort of females using the Registered Persons Database (RPDB) (Ontario) and the Consolidation File (BC) who were not pregnant during our timeframes of interest for Hypothesis 1.2.

#### Outcomes

We will use COVaxON in ICES and COVID-19 Immunization Data in PopData to define our COVID-19 vaccination outcome. Tdap vaccination status will be derived from the Ontario Laboratory Information System (OLIS); Tdap records are not available in PopData. OLIS and the MSP dataset in BC will define gestational diabetes screening outcomes. The Discharge Abstract Data (DAD) will provide data for our postpartum length-of-stay outcome. New onset of depression, anxiety or adjustment disorder will be ascertained using Ontario Health Insurance Plan (OHIP) diagnostic codes from physician billing claims, the DAD, and the Ontario Mental Health Reporting System (OMHRS) in Ontario; the BCPDR and physician billing claims report cases of depression, mood and anxiety. The timeframes of interest for our outcomes are illustrated in Figure 2.

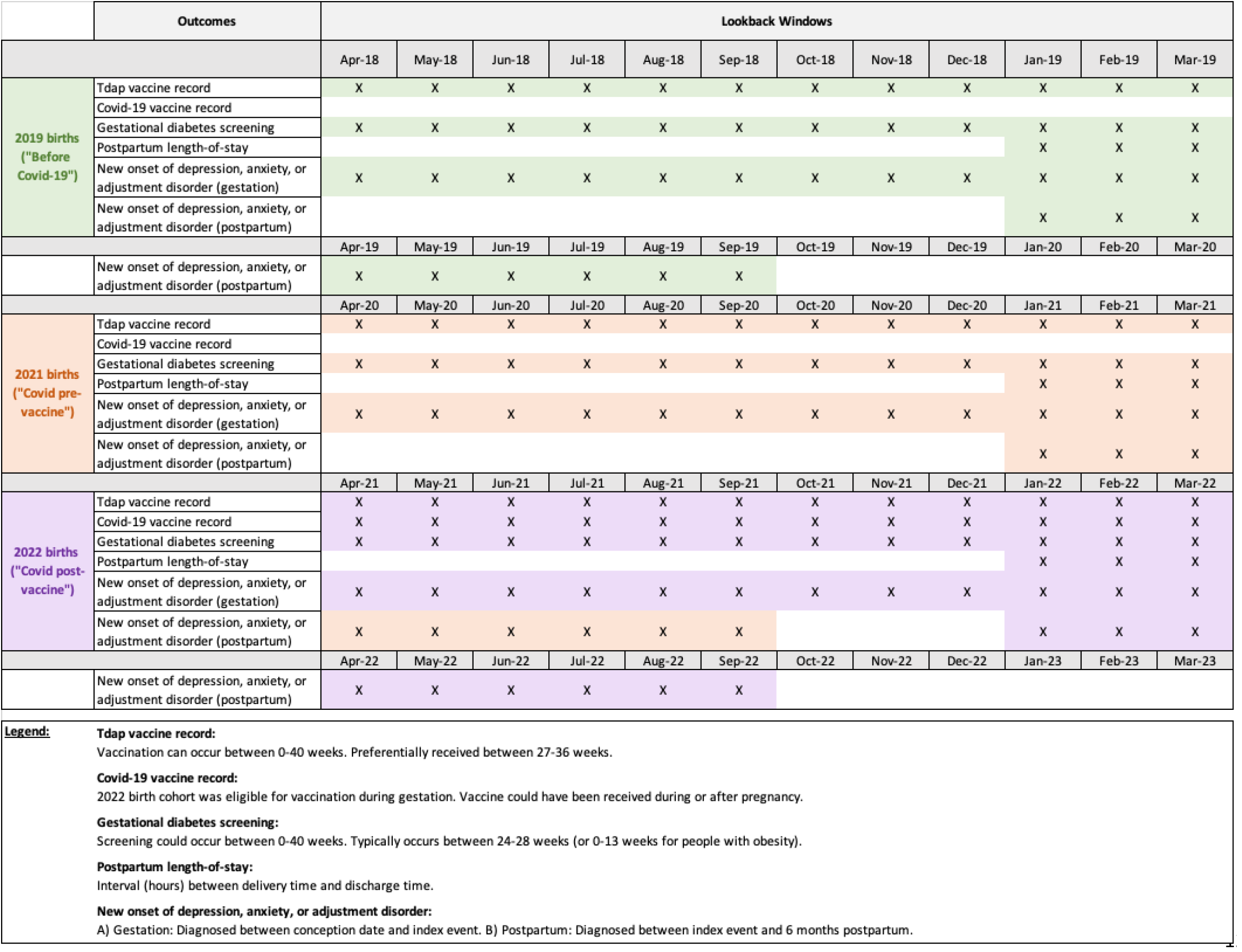

Data from the OHIP claims database and Medical Services Plan (MSP) dataset in BC will provide information about publicly funded perinatal services. The Registered Persons Database (RPDB) in Ontario, the BC Vital Events and Statistics Births dataset, the DAD, and the National Ambulatory Care Reporting System (NACRS) will provide information on baseline characteristics, covariates, and confounders of interest. The Canadian Index of Multiple Deprivation (CIMD) provides data on area-level socioeconomic and ethnocultural markers of marginalization.

#### Sample Size

Each year, there are approximately 140,000 live births in Ontario and 42,000 in BC.^59^ Our total population of January 1 to March 31 births over the three-year study period is, therefore, approximately 105,000 in Ontario and 31,500 in BC. Given our use of population-based health administrative data sources, we anticipate achieving large sample sizes to detect differences in outcome rates across groups of interest. Appendix 2 contains sample size calculations, based on anticipated incidence rates. When we report the findings from this study, we will compute post-hoc power calculations to demonstrate the adequacy of our sample in detecting differences. Guidelines for regression suggest assessing no more than 1 potential candidate predictor per 10-20 events.^60 61^

### Quantitative analyses

#### Descriptive

The primary goal of the quantitative analysis is to describe and compare rates of our outcomes across birth groups to help explain the qualitative findings. Pregnancy-related factors will be reported using measures of general frequency (i.e., counts and proportions), central tendency (i.e., means and medians), and dispersion (i.e., standard derivations and interquartile range). We will examine maternal age, rurality, CIMD measures (i.e., ethnocultural composition, economic dependency, and residential instability), medical practice characteristics of the mother’s perinatal care provider, parity, mental health history (e.g., previous diagnoses or emergency department visits for mental health), the use of assistive fertility, delivery characteristics (e.g., gestation length and delivery type), and newborn outcomes (e.g., infant birth weight and neonatal intensive care unit admission). These factors were identified *a priori* based on a review of the literature and consultations with theme group experts.

Data will be examined for completeness, the presence of outliers, and the nature of the variable distribution for continuous variables (e.g., skew). We will report the completeness range for each exposure variable. Cases with missing data will be deleted within each analysis. Where data are incomplete for key variables, we will consider using multiple imputation.

#### Inferential

The secondary analysis is to test for associations between our outcomes, primary exposure (birth group), and other covariates using multivariable logistic and linear regression. Covariates will be selected based on review of the literature, affirmed by topic and clinical experts. We will utilize logistic regression for our binary outcomes (i.e., vaccination status, gestational diabetes screening, and new onset of depression, anxiety, or adjustment disorder) and linear regression for our continuous outcome (i.e., postpartum length-of-stay). The best-subset approach will be used to identify the subset of covariates that best define the relationship with each outcome. This approach was selected to avoid suppressor effects commonly found with stepwise methods and statistically pre-determined predictors (e.g., *p* < 0.25). The Akaike information criterion (AIC) will be used to evaluate fit of the multivariable logistic regression models; R^2^ will be used to assess goodness of model fit for linear regression. We will verify model assumptions and employ methods such as robust variance estimation to address violations. We will also stratify these models by area-level information on socioeconomic status and ethnocultural composition to examine differences for low-income and racialized sub-groups.

All analyses will be performed using Statistical Analysis Software (SAS), version 9.4 (SAS Institute, Inc., Cary, North Carolina).

### Qualitative Design and Methods

This strand uses a qualitative descriptive approach ^62-64^ which is most suitable for integrating with quantative analytic outputs, since both rely on post-positivist epistemology. Qualitative description is a low-inference applied health research methodology which instructs analysts to stay close to the explicit meaning of participants, rather than progressing to abstraction or interpretation.

#### Participants

Eligible participants are people aged 18 or over who gave birth to a live baby in Ontario or BC between May 2020 and December 2021. Participants must be comfortable completing an interview in English.

#### Sampling and Recruitment

We will sample those who gave birth from May 1, 2020-Dec 1, 2021 to balance concerns of recall with information about different pandemic contexts. Guidance from one co-investigator with lived experience suggests that the complexity of health decision-making during the pandemic enhanced memory of this experience. We will use purposive sampling strategies, beginning with maximum variation to enroll an initial sample that represents diversity in age, SES, parity, race, geography, and occupational exposure to COVID-19. Preliminary analysis will suggest sampling strategies for future participants likely to yield analytically relevant findings. We will use a multi-pronged recruitment strategy, including posting advertisements online, in clinics and public parenting spaces. These advertisements will direct potential participants to an online or paper consent form which will explain the study and ask for socio-demographic information to be used for purposive sampling.^65^

#### Sample Size

We anticipate requiring 60-75 interview participants to reach sufficient information power that ensures our findings are credible and trustworthy.^66^ This sample will be split between Ontario (∼2/3) and BC (∼1/3) to reflect the population differences between the provinces. This determination was made based on the heterogeneity of the sample, the three-pronged aim of the study, the use of established theory and the anticipated medium quality of dialogue.^66^

#### Interview guide development

The interview guide was developed iteratively, with input from co-investigators who brought clinical, research, and lived experience of pandemic pregnancy. It was piloted and refined with three individuals who gave birth during the pandemic. The interview guide starts with a discussion of pandemic circumstances (family, housing, employment) and then elicits self-perceived risk of COVID-19 infection, complications, and long-term ramifications. Finally, the interviewer will inquire about decision-making processes relevant to the three topics of focus for this investigation (vaccination, prenatal care, mental health and social support).

#### Data Collection

Each enrolled participant will participate in one individual, semi-structured interview. The interviews will be conducted by telephone, Zoom, or in-person as participant preferences and geographical restrictions allow. Pilot data collection indicates that the interviews will last between 30-60 minutes. Interviews will be audio-recorded and transcribed verbatim.

### Qualitative analyses

Multiple analysts, including those with lived experience, will participate in conventional (inductive) content analysis,^67 68^ using a staged process of coding adapted from Grounded Theory.^69^ Analysis will focus on each theme separately, in conjunction with initial interview data about circumstances and risk perception. A multi-site qualitative analytic approach will maintain the site-specific context in the interviews while still offering comparative opportunity by theme.^70^ An audit trail will be maintained, comprising of memos from each stage of data collection and analysis. Findings will be triangulated between analysts, and participants. Resonance will be ensured by presenting emerging finding to a focus group of pregnant people.

### Data Integration

Data integration will occur at two points: during design and during interpretation. The project was designed with an objective framework-based mixing approach to develop complementary approaches for each of the three prenatal care themes.^71^ After independent data collection and analysis, we will integrate findings at the point of interpretation, using a joint display technique, to compare changes in health behaviours with qualitative data explaining those changes.^72^

### Strengths and Limitations

This study has several strengths. The large administrative datasets offer significant statistical power to investigate changes in health care use, stratified by markers of socioeconomic and ethnocultural status. The cross-provincial nature of our analysis allows the observation of consistency or change based on different healthcare policies, vaccine availability, medical care provision, and COVID-19 burden. The integration with qualitative data offers the potential for explaining observed differences. Our study team incorporates multiple individuals with lived experience of a pandemic pregnancy, and frontline clinicians who cared for pregnant people in both provinces during the pandemic.

This study has some limitations. Relying on administrative data hinders the ability to gather some data points of interest. For example, neither ICES or PopData have patient-level demographic data about race or socioeconomic status. Instead, we will rely on area-level data abstracted from the 2016 Canadian Census, operationalized through the CIMD. The administrative datasets exclude some births, such as those which took place outside of a hospital setting. Qualitative data will be gathered from people who gave birth between May 1, 2020 to Dec 1, 2021, which may introduce some challenges with recall and social desirability bias. However, our co-investigators with lived experience assure us that due to the unique and challenging nature of these decisions, they are able to recall their decision-making very clearly.

### Ethics & Dissemination

Ethics approval was received from McMaster University via the Hamilton Integrated Research Ethics Board for the qualitative strand (14632) and for the BC portion of the quantitative strand (15100C). Approval for the project was also received from the University of British Columbia Behavioural Research Ethics Board (H22-01905). Ethics approval was waived by McMaster University for the portion of the quantitative strand using ICES data (September 22, 2022) because the use of ICES data in this project is authorized under section 45 of Ontario’s Personal Health Information Protection Act (PHIPA) and does not require review by a Research Ethics Board.

This research will yield understanding about how a global pandemic and self-perceived risk influences health decision-making, which may inform health promotion initiatives through clinical counselling, public health communication and resource allocation, and policy development. Findings will be disseminated through peer-review manuscripts, at academic conferences, through clinical briefing notes and patient-facing materials.

## Data Availability

Qualitative data are not available for sharing when participants consented to participate in the study, they did not consent to access transcripts or data beyond what is reported within this report. The dataset for the quantitative portion of this study is held securely in coded form at ICES and PopData BC. While legal data sharing agreements between these data stewards and data providers prohibit ICES and PopData BC from making the dataset publicly available, access may be granted to those who meet pre-specified criteria for confidential access. Readers are welcome to contact the research team for further information.

## Protocol

Available to interested readers by contacting Dr. Vanstone

## Computer code

The full dataset creation plan and underlying analytic code are available from the authors upon reasonable request, understanding that the computer programs may rely upon coding templates or macros that are unique to ICES and are therefore either inaccessible or may require modification.

## Author contribution

MV, DG, ED, MH, HB, AD, JL, TP, MP, MM, SK, SDM contributed towards the research idea and provided input into the design of the study. RC, KS, CK, AC, MP, SK, MM, MV, DH, EH, MS, NS, CM, AJ participated in the design of qualitative or quantitative data collection instruments and analytic plan. MV and RC drafted and revised the manuscript. All authors provided critical feedback and approved the version to be published.

## Data

Qualitative data are not available for sharing; when participants consented to participate in the study, they did not consent to access transcripts or data beyond what is reported within this report. The dataset for the quantitative portion of this study is held securely in coded form at ICES. While legal data sharing agreements between ICES and data providers prohibit ICES from making the dataset publicly available, access may be granted to those who meet pre-specified criteria for confidential access, available at www.ices.on.ca/DAS. Readers are welcome to contact the research team for further information.

